# Hyperbaric Oxygen Therapy for Long COVID: 3-Month Follow up Results from a Prospective Registry of 232 patients

**DOI:** 10.1101/2024.09.02.24312948

**Authors:** J. van Berkel, R.C Lalieu, D. Joseph, M. Hellemons, C.A. Lansdorp

## Abstract

A potential beneficial effect of hyperbaric oxygen therapy (HBOT) on complaints of long COVID was found, leading to increased demand for this treatment despite many remaining clinical questions and lack of formal guideline recommendations and reimbursement. A registry was set up in order to gain more insight into patient characteristics and (long-term) outcomes of long COVID patients undergoing HBOT. Patient-reported outcome measures were collected at baseline, after treatment and at 3-month follow up. The primary outcome measures were the mental and physical component score (MCS/PCS) of the SF-36 questionnaire 3 months after HBOT. A clinically relevant positive or negative response was defined as an increase or decrease of ≥10% in MCS and/or PCS after 3 months. Secondary outcomes included the EQ-5D, severity of complaints and ability to work. In this prospective registry of 232 long COVID patients, 65% of long term-ill patients had a clinically relevant increase in quality of life. However, 15% of the patients experienced deterioration in quality of life. Symptoms that showed most improvement were predominantly in the cognitive domain. This indicates that HBOT may have a positive effect on complaints of long COVID, but alertness for worsening of the condition should be exercised.

## INTRODUCTION

Although the majority of people recover fully from an infection with SARS-CoV-2, 6-12% of the adults develops longer lasting symptoms.^[1,2]^ This condition, referred to as long COVID (or post COVID), was recently defined as: *an infection-associated chronic condition that occurs after SARS-CoV-2 infection and is present for at least 3 months as a continuous, relapsing and remitting, or progressive disease state that affects one or more organ systems*.^[4]^ A wide range of symptoms can occur, with the most commonly reported symptoms: fatigue, post-exertional malaise and cognitive problems. Frequently reported cognitive symptoms include memory and concentration problems, difficulty processing stimuli and brain fog.^[3,4,5,6]^ The condition can last for months or years or become a chronic condition.^[3,5,6,7]^

Unsurprisingly, the impact of long COVID is substantial and leads to decreased quality of life, social functioning and has a major impact on work life.^[2,3,4,5,6]^ In the Netherlands, 18% of long COVID patients reported being unable to work and 41% had to significantly reduce their working hours.^[6]^

Research on long COVID has known a slow and difficult beginning, and there is still incomplete understanding of its complex pathophysiology. Proposed mechanisms include immune-dysregulation, microbiome disruption, autoimmunity, clotting and endothelial abnormalities, mitochondrial dysfunction, viral persistence or dysfunctional neurological signaling.^[2]^ Corresponding with the lack of understanding, treatment options for long COVID are limited; common approaches include coping strategies, activity management and symptom control.^[3,5]^

In search of an effective treatment, hyperbaric oxygen therapy (HBOT) emerged as a promising intervention. HBOT is a medical treatment where an individual breathes near 100% oxygen intermittently while inside a hyperbaric chamber that is pressurized to greater than 2 atmosphere absolute (ATA).^[8]^ HBOT is used for established indications such as diabetic foot ulcers, late radiation tissue injury and emergency indications like decompression illness.^[8]^ HBOT increases tissue oxygen levels and activates genes that trigger regenerative processes, including stem-cell proliferation, anti-inflammatory responses, angiogenesis, and can benefit mitochondrial function.^[9,10]^ Furthermore, it influences cerebral blood flow and microstructure, aiding neuroplasticity and cognitive function.^[11,12]^ Previous studies suggest improvement of long COVID symptoms after HBOT.^[13,14,15,16,17,18,19,20,21,22]^ A sham-controlled, randomized trial showed significant improvements in cognitive symptoms, quality of life, sleep, psychological symptoms and pain after 40 sessions of HBOT.^[15]^ Longitudinal follow up of HBOT patients in this trial showed persistent improvement in patient-reported outcomes one year after treatment.^[16]^ Nonetheless, although effectiveness was demonstrated, the clinical relevance of its effects was questioned, and its efficacy and safety remains to be established in the more severe and longer term-ill patients, in whom spontaneous recovery is less likely.

Based on the existing studies, the Association of Hyperbaric Medicine in the Netherlands decided the current evidence allows further exploration of HBOT for long COVID patients.^[23]^ In order to gain more insight into outcomes of this treatment, patients who decided to undergo (off-label) HBOT for long COVID were invited to participate in a prospective registry. In this article the 3-month follow up results from this registry are presented.

## MATERIALS AND METHODS

### Study design and patient selection

Six out of nine hyperbaric chambers in the Netherlands participated in this prospective registry. All consecutive patients that were deemed eligible for treatment between 15 February 2023 and 15 February 2024 were asked to participate in the registry and written informed consent for the acquisition of data and patient reported outcome measures (PROMS) was obtained prior to the start of treatment. Patients who were not able to provide consent or fill in questionnaires were excluded, as well as patients that were eligible for treatment but did not start the treatment. The study was approved by the Medical Ethical Review Board of Erasmus University of Rotterdam (MEC-2023-0433), and was documented in a trial registry (clinicaltrials.gov, NCT06159309).

Local eligibility criteria for referral and off-label treatment in these centers were in line with previous literature: age ≥ 18 years, (primary) cognitive complaints with significant impact on everyday life for at least 3 months after initial COVID infection and with no alternative explanations for these symptoms. Initial COVID infection was preferably confirmed by PCR or antigen test or was highly probably (e.g. patients with typical COVID symptoms during the endemic phase before testing was possible). Eligibility for the treatment and contraindications were assessed by the referring specialist and the hyperbaric physician.

### HBO regimen

The local protocol for off-label treatment comprised 40 daily sessions of HBOT at 2.4-2.5 ATA, with a treatment duration of 90 to 110 minutes with 5-minute air breaks every 20 minutes. Treatment was provided 5 days a week (excluding weekends), with a total treatment duration of 8 weeks (in the case of 40 sessions). This protocol was based on prior studies for this indication and other national protocols for established indications.^[13,14,15,16,17,18,19,20,21,22]^

At the discretion of the hyperbaric physician extension of the protocol was possible. Reasons for prolonging the treatment were either clinical (e.g. a subjective late response with the wish to continue treatment at the moment of response) or because of inadequate dosing in the beginning of treatment due to interruptions of the treatment schedule (e.g. due to illness).

Regular visits with the hyperbaric physician and/or case manager were scheduled at baseline and after every 10 to 15 sessions and in the case of extension of treatment also after 50 or 60 sessions, in accordance with standard care at the facilities. During these visits the progression of the treatment and complaints were discussed, as well as any adverse events. Follow up was performed by telephone: the 3-month follow up is part of standard care of HBOT.

### Outcome measures

Patients were asked to fill out PROMS at baseline, after 40 sessions and 3 months after treatment. In the case of extension of treatment beyond 40 sessions, PROMs were also collected directly after the extended treatment (i.e. after 50 or 60 sessions).

The primary endpoint is the mental and physical component score (MCS/PCS) of the SF-36 questionnaire. Each component score measures quality-of-life on four health domains: MCS includes role limitations due to emotional problems, energy/fatigue, emotional well-being and social functioning. PCS includes general health, physical functioning, role limitations due to physical health and pain. Scores range from 0 (lowest) to 100 (highest). No minimally clinical important difference (MCID) for the SF-36 is available in literature for long COVID patients specifically, and therefore a 10% increase in score or more is used in this study as MCID, as is in line with the national guideline.^**[24]**^ Responders after HBOT were defined as ≥10% increase in MCS and/or PCS score between baseline and 3-month follow up. Patients who had a ≥10 deterioration of MCS and/or PCS score were defined as negative-responders. The patients who had <10% change of MCS and PCS or a mixed result (e.g. positive response in MCS, but negative in PCS) were defined as non-responders.

Secondary endpoints include:

▪ EQ-5D questionnaire: The EQ-5D evaluates quality of life in 5 domains (mobility, self-care, usual activities, pain/discomfort and anxiety/depression) and a visual analogue scale (VAS), which measures quality of health on a scale ranging from 0 (worst imaginable health) to 100 (best imaginable health). A MCID of 7.5 for the VAS has been reported for patients with long COVID.^[25]^
▪ Questionnaire of long COVD symptoms: patients evaluated the presence and severity of 40 common long COVID symptoms. The list of symptoms was based on available literature. Patients rated the presence and severity of their symptoms on a 5-point Likert scale categories (not present/mild/moderate/severe/extreme) and were then further subcategorized into: no symptoms, mild-moderate and severe-extreme.
▪ Absence from work: all patients were questioned regarding absenteeism from work and its relation to their diagnosis of long COVID, number of working hours and adjustments on the job content.
Furthermore, information on adverse events of HBOT (both solicited during regular visits and unsolicited), re-infection and re-vaccination during HBOT and follow up was recorded.

### Data management and statistical analysis

An external electronic data capture system (Castor EDC) was used to document and store patient information between centers. Baseline characteristics were recorded during the initial assessment by the hyperbaric physician (before the start of treatment), and included gender, age, patient body mass index, smoking, medical history, and COVID specific information including the date of infection leading to long COVID, hospital or intensive care admission, type of test (PCR or antigen) and COVID vaccination history.

IBM SPSS Statistics 29.0.1.0 software was used to analyze all data. Descriptive statistics were used to describe patient baseline characteristics. Categorical data are presented as number and/or percentage, and continuous data are presented as median with an interquartile range (IQR). For evaluation of outcomes, distribution of the data was assessed on histograms, if normality was assumed this was tested (Kolgorov-Smirnov). To compare groups, paired T-tests were used if data was normally distributed, Wilcoxon signed rank tests were used if data was not normally distributed. A value of p<0.05 was considered statistically significant.

## RESULTS

### Registry

Between February 2023 and February 2024, 239 patients were eligible for inclusion. 5 patients were excluded because they never started HBOT due to private circumstances, and 2 patients did not provide informed consent. Therefore, 232 patients were included in the final analysis. Of the 232 patients, 187 patients were treated at Eurocept Clinics (Amersfoort, Arnhem, Geldrop, Hoogeveen, Rotterdam and Waalwijk) and 45 patients at the Hyperbaric Medical Center in Rijswijk. The response rate on the primary outcome (SF-36) was 95% at baseline, 84% directly after HBO and 80% at 3-month follow up.

### Baseline characteristics

Baseline characteristics are presented in Table 1. The median age of patients at the start of the treatment was 42 years (IQR 31 -51). Most patients were female (70%) and never-smokers (76%), with only 2% patients being active smokers at the start of treatment. 118 patients had no documented medical history. Of the reported history items in the remaining 114 patients, mental complaints (12%), asthma (11%), irritable bowel syndrome (7,3%), hypertension (4,7%) and other cardiovascular disease (4,7%) were most frequently reported. Most COVID infections that lead to long COVID were confirmed by PCR (71%) or antigen testing (15%). A minority of patients (3%) had been hospitalized for their COVID infection, with none being admitted to the intensive care unit. Patients had long-term complaints before starting HBOT with a median 20 months (IQR 14 -30) between SARS-CoV-2 infection and start of HBOT.

**Table 1.** Patient characteristics (N=232)

|  |  |  |
| --- | --- | --- |
| Age |  | 42 (31 - 51) |
| Body Mass Index<sup>i</sup> |  | 24,2 (21,9 - 27,1) |
| Sex | Female | 162 (70%) |
|  | Male | 70 (30%) |
| Smoking history | Smoker | 5 (2%) |
|  | Ex-smoker | 51 (22%) |
|  | Never-smoker | 175 (76%) |
| COVID test | PCR+ | 156 (71%) |
|  | Antigen+ | 33 (15%) |
|  | Not confirmed by test (first wave) | 31 (14%) |
| Hospital admission |  | 6 (3%) |
| - Intensive care admission |  | - 0 (0%) |
| Months between SARS-CoV-2 infection and start of treatment |  | 20 (14 - 30) |
| COVID vaccinated (received at least one) |  | 207 (95%) |
| Medical history |  | 114 (49%) |
| - Mental disorders |  | 27 (11,6%) |
| - Asthma |  | 26 (11%) |
| - Irritable bowel disease |  | 17 (7,3%) |
| - Hypertension |  | 11 (4,7%) |
| - Other cardiovascular disease |  | 11 (4,7%) |
| - Auto-immune disease |  | 10 (4,3%) |
| - Infectious mononucleosis |  | 8 (3,4%) |
| - ADHD / ADD |  | 7 (3,0%) |
| - Other gastrointestinal disease |  | 7 (3,0) |
| - Neurological/neuromuscular disorder |  | 7 (3,0%) |
| - Burn-out |  | 7 (3,0%) |
| - Thyroid disease |  | 3 (1,3%) |
| - Other pulmonary disease |  | 3 (1,3%) |
| - Intestinal parasitic infection |  | 2 (0,9%) |
| - Chronic kidney disease |  | 2 (0,9%) |
| - Diabetes |  | 1 (0,4%) |
| - CFS/ME |  | 1 (0,4%) |
| - Immune deficiency |  | 1 (0,4%) |
| - Other |  | 19 (8,2%) |

Numbers are shown as *n* (%). Continuous variables are shown as median (IQR).
Abbreviations: *IQR* interquartile range. ADHD/ADD, Attention Deficit (Hyperactivity) Disorder. CFS/ME, chronic fatigue syndrome/myalgic encephalomyelitis.
<sup>i</sup> Calculated as weight/height<sup>2</sup>

### HBOT sessions

Patients were treated with a median of 40 sessions (IQR 40 -46). 15 (6%) patients terminated their treatment before 37 sessions. Reasons for early termination of the treatment were: HBOT was too exhausting with worsening of fatigue or other long COVID symptoms (10 patients), anxiety or claustrophobia during HBOT (2 patients), no treatment effect (1 patient), an unrelated illness (1 patient), and private circumstances (1 patient).

54 patients received 48 sessions or more. 42 patients because of a (subjective) late response with the wish to extend the treatment, and 12 patients because of inadequate dosing in the beginning of treatment due to interruptions of the treatment schedule (e.g. due to illness).

### Primary outcome: SF-36 questionnaire

The median MCS of the SF-36 questionnaire improved from 45,5 (IQR 28,7 -56,6) at baseline to 57,0 (IQR 46,2 -71,5) at 3-month follow up. This improvement was statistically significant (p<0,001). For the PCS, these scores were 36,9 (IQR 27,2 -45,0), and 42,5 (IQR 32,2 -52,5); p<0,001, respectively. Subdomains that showed the largest increase in score were social functioning, energy/fatigue and physical functioning (see figures 1a and 1b for median scores, and additional information in table A in the appendix).

**Figure 1a.**
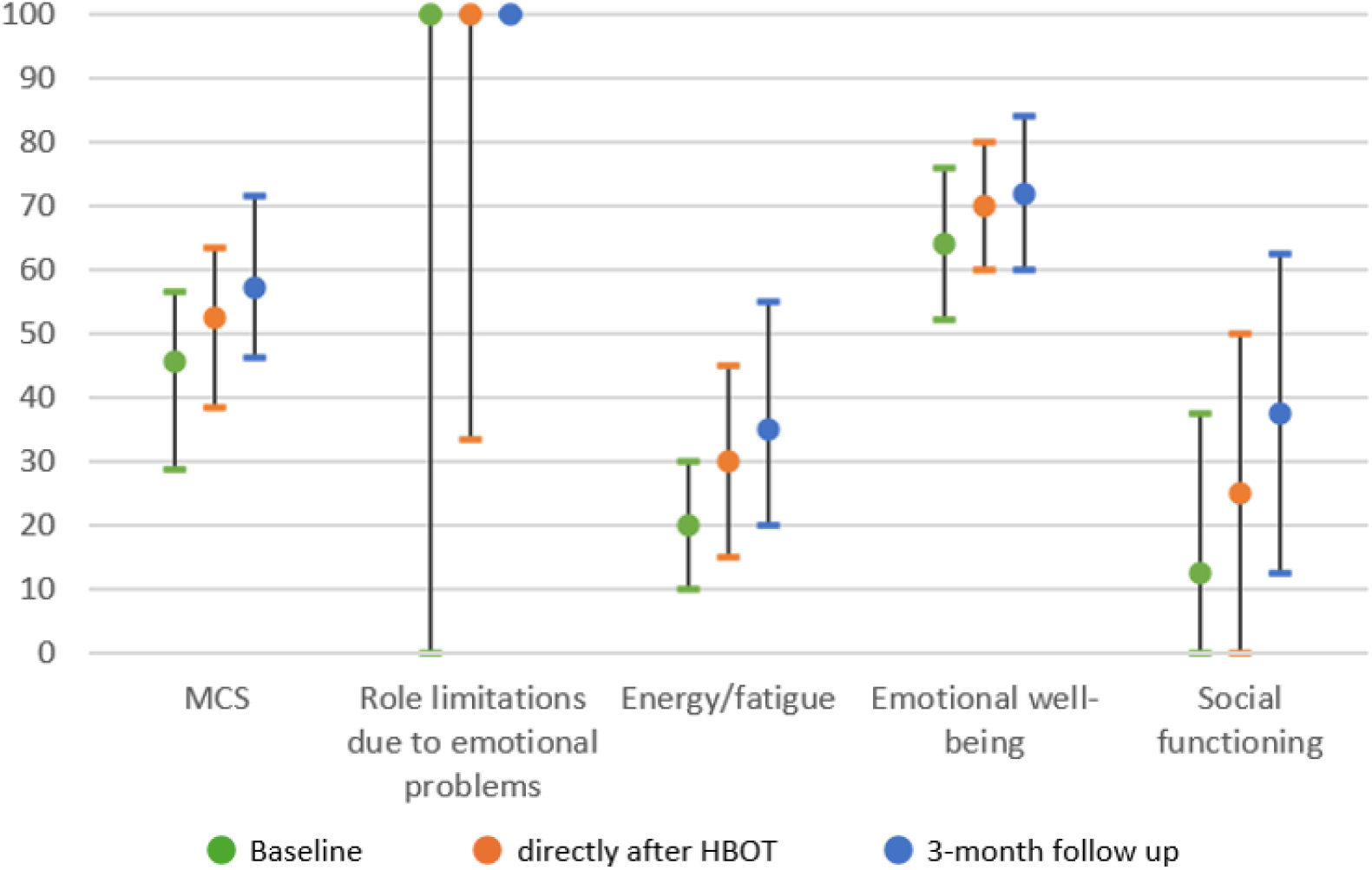
SF-36 Mental component scores.

**Figure 1b.**
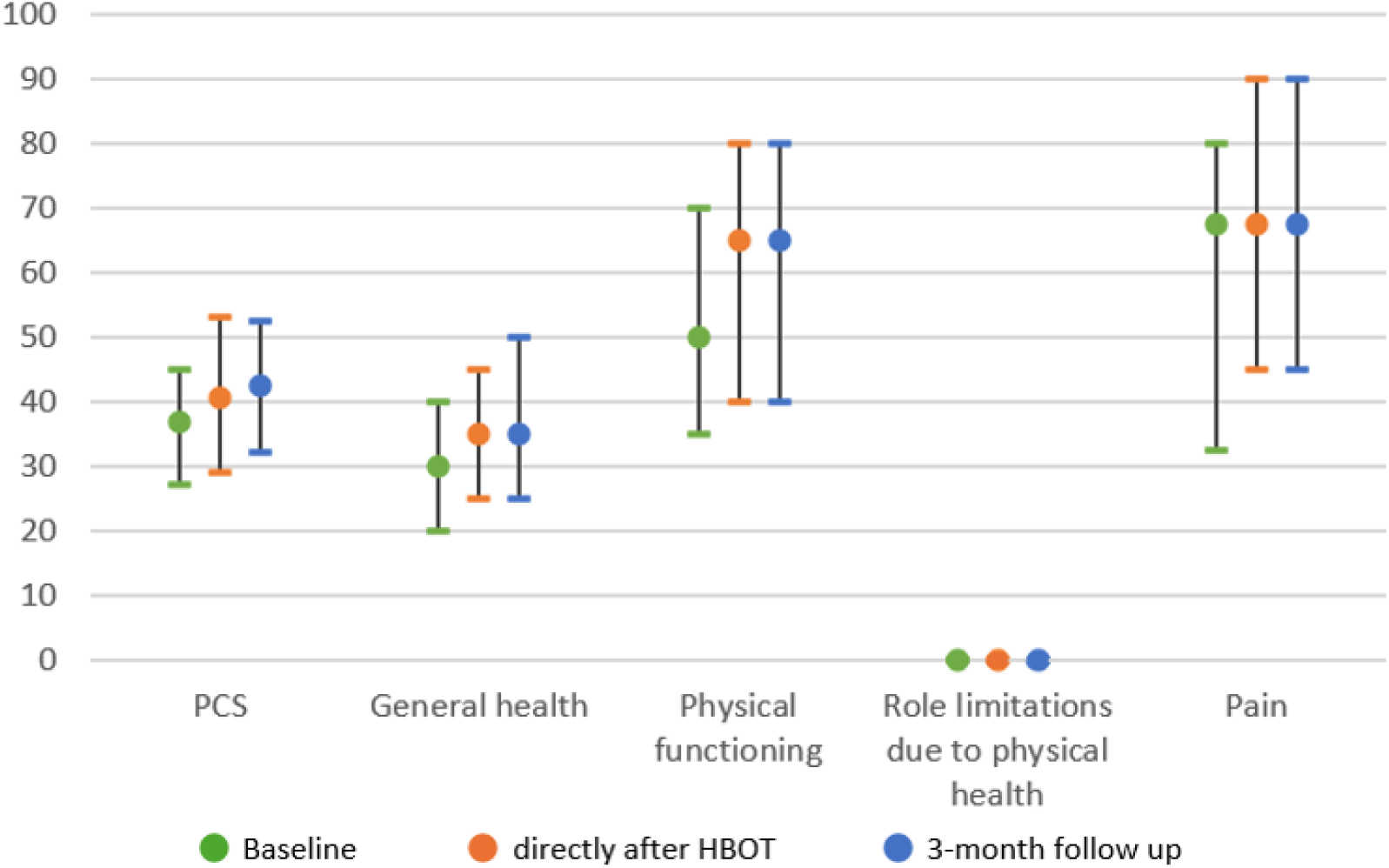
SF-36 Median physical component scores. Note for Figure 1a/b:The SF-36 measures quality-of-life. The MCS of the SF-36 measures quality-of-life on four health domains: role limitations due to emotional problems, energy/fatigue, emotional well-being and social functioning. The PCS includes general health, physical functioning, role limitations due to physical health and pain. Scores range from 0 to 100, with higher scores indicating higher quality of life. Median scores are displayed as dots, with an interquartile ranges as lines, corresponding values can be found in Table A in the appendix.Abbreviations: SF-36, 36-item Short Form Survey. PCS, physical component score. MCS, mental componentscore.

3 months after HBOT, out of 138 patients, 90 (65%) patients were responders, 27 (20%) were non-responder and 21 (15%) were negative-responders. Patient characteristics for each subgroup are shown in Table 2.

**Table 2.** Responder, non-responder and negative-responder patient characteristics.

|  |  | Responder<br>(N=90) | Non-Responder<br>(N=27) | Negative-Responder (N=21) |
| --- | --- | --- | --- | --- |
| Age |  | 43 (33 -52) | 39 (28 - 51) | 42 (37 - 55) |
| Body Mass Index<sup>i</sup> |  | 24,7 (22,7 - 27,5) | 22,6 (19,7 - 24,4) | 24,0 (21,1 - 27,2) |
| Sex | Female | 54 (60%) | 22 (81%) | 17 (81 %) |
|  | Male | 36 (40%) | 5 (19%) | 4 (19%) |
| Smoking history | Smoker | 1 (1%) | 0 (0%) | 0 (0%) |
|  | Ex-smoker | 21 (24%) | 5 (19%) | 7 (33%) |
|  | Never smoker | 67 (75%) | 22 (81%) | 14 (67%) |
| COVID test | PCR+ | 59 (70%) | 19 (73%) | 13 (62%) |
|  | Antigen+ | 13 (14%) | 3 (12%) | 4 (19%) |
|  | Not confirmed by test (first wave) | 13 (14%) | 4 (15%) | 5 (20%) |
| Hospital admission |  | 3 (3%) | 0 (0%) | 0 (0%) |
| Months between SARS-CoV-2 infection and start of treatment |  | 19 (13 - 30)) | 18 (14 - 26) | 20 (14 - 31) |
| COVID vaccinated (at least one) |  | 78 (92%) | 25 (96%) | 19 (95%) |
| Medical history, yes/no |  | 42 (47%) / 48 (53%) | 16 (59%) / 11 (41%) | 11 (52%) / 10 (48%) |
| SF-36 component scores<sup>ii</sup>: |  |  |  |  |
| - MCS at baseline |  | 41,2 (23,3 - 56,7) | 46,4 (37,5 - 61,9) | 54,3 (47,2 - 59,3) |
| - PCS at baseline |  | 35,3 (26,1 - 45,0) | 37,5 (29,4 - 47,5) | 35,6 (22,5 - 55,0) |
| EQ-5D VAS score<sup>iii</sup> |  | 40 (30 - 50) | 40 (29 - 50) | 35 (27 - 40) |
| Re-infection with SARS-CoV-2 during or after HBOT |  | 9 (10%) | 3 (11%) | 2 (10%) |
| Re-vaccination (COVID) during or after HBOT |  | 7 (8%) | 2 (7%) | 2 (10%) |

Note: Numbers are shown as *n* (%). Continuous variables are shown as median (IQR).
Abbreviations: *IQR* interquartile range. ADHD/ADD, Attention Deficit (Hyperactivity) Disorder. CFS/ME, chronic fatigue syndrome/myalgic encephalomyelitis. SF-36, 36-item Short Form Survey. PCS, physical component score. MCS, mental component score. VAS, visual analogue scale. HBOT, hyperbaric oxygen therapy.
<sup>i</sup> Calculated as weight/height<sup>2</sup>
<sup>ii</sup> The SF-36 measures quality-of-life. The MCS of the SF-36 measures quality-of-life on four health domains: role limitations due to emotional problems, energy/fatigue, emotional well-being and social functioning. The PCS includes general health, physical functioning, role limitations due to physical health and pain. Scores range from 0 to 100, with higher scores indicating higher quality of life.
<sup>iii</sup> The EQ-5D VAS score records the respondent's self-rated health on a visual analogue scale from 0 to 100, with higher scores indicating higher quality of life.

In patients who received an extension of their treatment beyond 40 sessions no difference was found in median MCS /PCS score after 40 sessions compared to after 50 or 60 sessions. Median MCS was 49,0 (IQR 32,5 – 55,4) after 40 sessions, and 48,1 (IQR 38,4 – 53,9) at 50 or 60 sessions, p=0,773, N=25. The median PCS score was 36,9 (IQR 21,6 – 43,4), and 33,8 (IQR 23,4 -43,4, p = 0,887, N =21), respectively. At 3-month follow up 73% of the patients who received extension of treatment to 50 or 60 sessions were responders and 21% were negative responders. In the patients who received 40 sessions this was 63% and 14%, respectively.

### EQ-5D

The mean score on the visual analogue scale of the EQ-5D improved from 37 (SD 14) at baseline to 50 (SD 19) at 3-month follow up (p<0,001). Out of 163 patients, 99 (61%) improved more than the MCID at 3-month follow-up, 16 (10%) patients had a deterioration of more than the MCID, and 48 patients (29%) did not improve or deteriorate more than the MCID.

### Long COVID complaints questionnaire

Out of the 40 included complaints in the questionnaire, the complaints that were most often scored ‘severe to extreme’ at baseline were fatigue (87%), difficulty processing stimuli (80%), post-exertional malaise (75%), loss of physical fitness (71%) and concentration problems (68%).

At 3-month follow up, for the top 5 symptoms there was a global decline in symptom burden, for example from severe-extreme symptoms to mild-moderate (figure 3). For fatigue 55/155 (35%) patients with severe-extreme symptom score had less severe complaints at 3 month follow up. On the other hand, 9/21 (43%) patients with no or mild-moderate symptoms transitioned to severe-extreme complaints at 3-month follow up. Respectively, for difficulty processing stimuli 81/146 (55%) reported improvement and 6/33 (18%) deterioration, for post-exertional malaise 45/136 (33%) reported improvement and 6/44 (14%) deterioration, for loss of physical fitness 42/123 (34%) reported improvement and 15/53 (28%) deterioration and for concentration problems 77/126 (61%) reported improvement and 5/54 (9%) deterioration.

**Figure 3.**
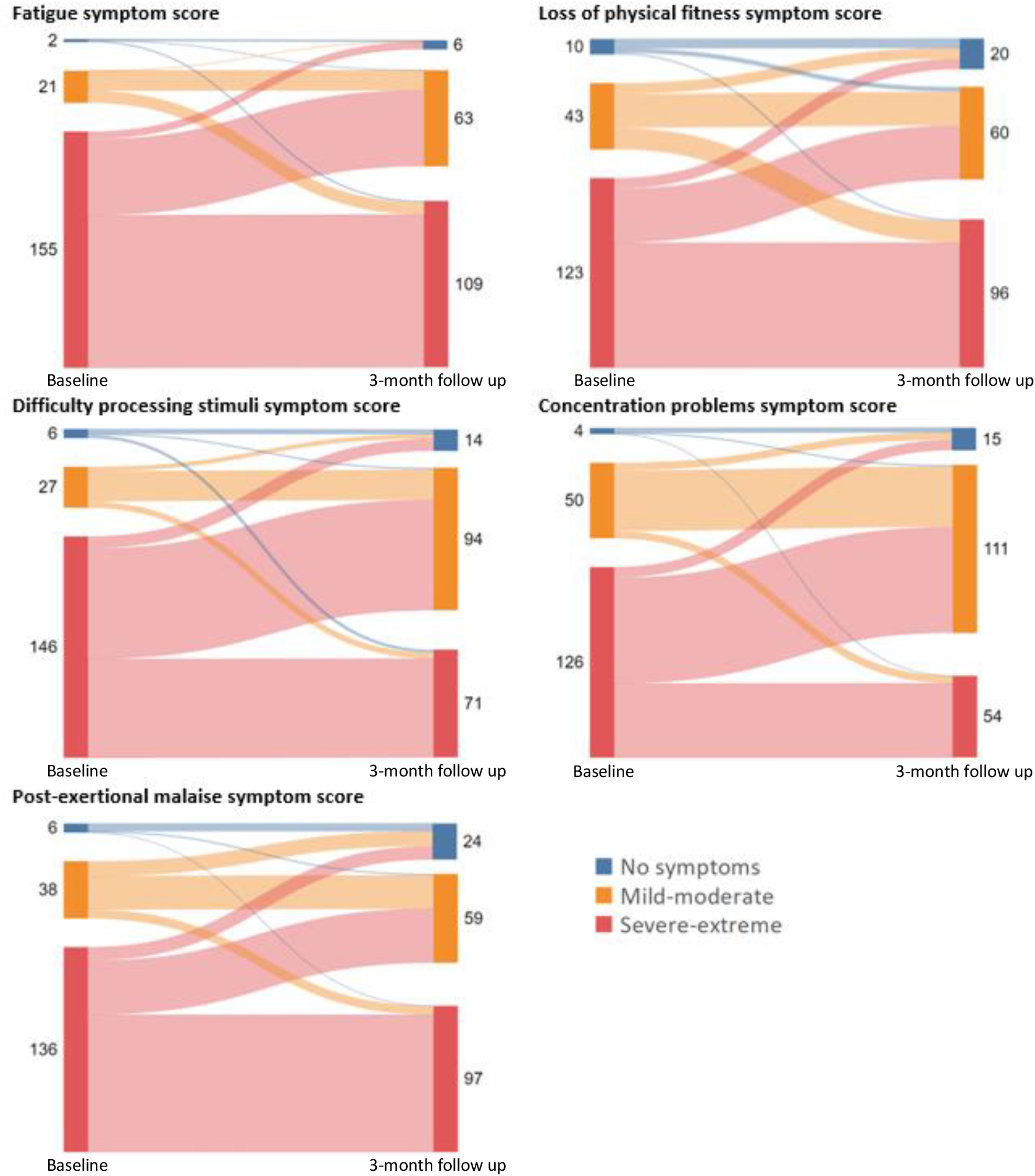
Severity of top-5 long COVID complaints at baseline and 3-month follow up. Note: Number of patients for each symptom category. Symptom severity was rated by patients on a 5-point Likert scale categories, and further subcategorized into; no symptoms, mild-moderate and severe-extreme.

The most substantial decreases in symptom burden (from severe-extreme to less severe) were seen for word-finding problems (43/59 patients, 73%), irritability (44/61, 72%), memory problems (62/88,70%), brain fog (70/103,68%) and shortness of breath on exertion (38/68,56%). All symptoms and frequencies over time are presented in supplemental table A and B.

### Absenteeism from work

In the registry there were 196 people who were working before long COVID complaints. At baseline, 91 (46%) of them were completely unable to work due to complaints of long COVID. Of the patients who were still able to work, 96% had reduced their weekly working hours to a median of 12 (IQR 8 -23) hours per week or 34% (IQR 22 - 60) of the hours stated in their contract. Furthermore 78 (74%) working patients reported adjustments in the content of their work because of their illness.

At 3-month follow up, 11% of patients who were not working at baseline due to long COVID were able to resume activities at work. On average, they were able to work for 9 (IQR 4 - 32) hours per week. Patients who were already working at baseline but who had reduced their working hours because of long COVID, were not able to work more hours per week at 3-month follow up (median 12 (IQR 8 - 20), p 0,294). Content of work was no longer adjusted due to complaints in 9% of patients.

### Adverse events

In the 232 patients who received HBOT there were 24 unsolicited adverse events. The most common unsolicited adverse event was barotrauma of the middle ear (11 patients). Furthermore, 2 patients had complaints of anxiety/claustrophobia during HBOT, and 1 patient had an episode with symptoms of shortness of breath. And, as stated before, 10 patients experienced worsening of fatigue or other long COVID symptoms during HBOT and terminated treatment before completion.

Solicited events during regular visits were fatigue (65%) and blurry vision (27%), which are common complaints associated with HBOT. These complaints were reversed at 3-month follow up, except for one patient who still experienced blurry vision. A consultation at the ophthalmologist revealed an incipient cataract, not requiring treatment at that time.

## DISCUSSION

Results from this prospective registry demonstrate that 61-65% of long COVID patients respond positively to HBOT in terms of quality of life, and response is maintained at least up to 3 months after treatment. The largest improvement was seen on the mental components of quality of life, including social functioning, energy/fatigue, and physical functioning. Cognitive symptoms improved in up to 73% of the patients previously reporting severe to extreme complaints in this domain at baseline. A minority of patients had already made steps in reintegration at work at 3-month follow up.

At the same time, a deterioration long COVID complaints was found in 10-15% of patients. Worsening of long COVID after HBOT was not reported in earlier research but represents an important safety signal. Given the high prevalence of post-exertional malaise and heavy symptom burden, HBOT treatment may be too exhausting for certain patients in this specific patient group, worsening their condition. The deterioration of long COVID symptoms during/after HBOT have led to adaptations in local protocols for the treatment: a minimum 4 hours of upright activity per day is now a criterion to be eligible for treatment as a proxy for sufficient resilience. Furthermore, patients are informed that an increase in fatigue, malaise or other complaints during HBOT can be a reason for early termination. Regular visits with the hyperbaric physician during the treatment, e.g. every 5 or 10 sessions, can help monitor any changes in the course of the disease.

Other adverse events were mostly minor and reversible. Nonetheless, it should be noted that one patient was diagnosed with incipient cataracts, after complaints of blurry vision did not improve in the months after HBOT. The patient did not have problems with visual acuity before the start of HBOT. Given the nature of this study a causal effect between HBOT and the occurrence of cataracts cannot be established nor ruled out. Induction of cataracts after HBOT has been described in literature but is associated with treatment schedules of >100 sessions. However, in a recent overview of the literature from 2024, unpublished results of an ongoing randomized HBOT trial are reported that show development of cataracts in approximately 2% of the patients receiving 20 to 40 sessions (n=120).^[26]^ This implies that the incidence of cataracts may be underestimated. For future research it is advised to inform patients about this possible side effect as part of the informed consent procedure. Furthermore, early referral for further examination should be warranted in case complaints of blurry vision do not show improvement within 10 to 12 weeks after treatment.

Local protocol at the hyperbaric chambers was a treatment schedule of 40 sessions, in line with the study that provided the highest level of evidence that is available for this indication as of now.^[15]^ Individual changes in this schedule were made at the discretion of the hyperbaric physician and/or referring physician, which happened in 23% of cases, with an addition of 10 or 20 sessions being most common. The percentage of responders (73%) was slightly higher in the group of patients who received an extended treatment compared to the patients who received 40 sessions (63%). However, the percentage of negative-responders was also higher in the group with extended treatment (21% versus 14%). Dose-response relationship is an interesting topic for future research, since the burden for patients and the cost of treatment is affected by more or less sessions. Other studies have suggested treatment protocols with fewer sessions could also be effective.^[13,19,21]^

The findings in this study add on to previously reported results but are unique with regards to the number of patients included and the longer-term follow up of 3 months. Furthermore, it provides real-life insight into the current situation in the Netherlands and the type and number of patients who opt for off-label therapy when made available. Moreover, the included patients are patients with more severe symptoms of longer duration that the earlier studies, who are less likely to recover spontaneously.

## STRENGTHS

A strength of the current study is that it took place in a real-life, clinical setting, with a diverse group of patients. We feel the current study functions as a proof of concept for successfully treating long COVID symptoms with HBOT in patients who are able to undergo the therapy. Since the infrastructure to provide this therapy is already in place, there are no concerns for the applicability of this treatment in clinical practice. Furthermore, we have used multiple questionnaires to measure improvement in quality of life in different domains, and symptoms specifically related to long COVID. The results are maintained for at least three months, and follow up one year after treatment is planned to investigate long-term results as well.

## LIMITATIONS

Limitations of this study are the lack of a control group, possible selection and confirmation biases and the questionnaires that were used. Given the uncontrolled nature of this study, a direct causal effect of HBOT on the outcomes cannot be established. For this a randomized, controlled trial is planned in the Netherlands, and data of this registry will serve as pilot-data for this future study. A control group would have served to notice any improvement due to time passing. However, the median time from COVID infection to the start of HBOT was 20 months, which makes natural resolution of symptoms less likely; studies suggests that recovery of long COVID appears to plateau after 6-12 months after infection, and few patients achieve full remission.^[5,6]^

Furthermore, the population in this study is comprised of patients who were able to 1) undergo the therapy and 2) finance their own treatment, since it is currently applied off-label. This has undoubtedly led to a selection bias of patients with enough means and capacities for this treatment, and who are highly motivated to complete the treatment. This motivation could also lead to confirmation bias as any improvement in symptoms might get attributed to the treatment. We feel this effect is limited, since there is still a substantial amount of non- or negative responders in the population. Finally, in this study, standard of care patient-reported outcomes (i.e. SF-36 and EQ-5D) were used. On top of this, a specific long COVID complaints questionnaire was added, as well as questions regarding work history. Although this set gives an important impression of the disease burden and improvement over time, a more comprehensive assessment would be of interest, for example more specifically focused on other specific disease aspects of long COVID such as post-exertional malaise burden and dysautonomia features. Recently, an international core outcome set for long COVID was proposed.^[27]^ For future prospective studies we aim to take such disease specific outcomes and features into account.

In conclusion, the majority of patients who opted for HBOT reported a significant and clinically relevant increase in quality of life 3 months after treatment. At the same time, a minority of patients deteriorated during or after HBOT. Often reported complaints, predominantly in the cognitive domain, responded positively to treatment. A small percentage of patients previously absent through sickness were able to return to work 3 months after treatment. These findings add onto previous research showing the potential of HBOT in long COVID in improving symptoms in moderate-severely affected and long-term ill patients with long COVID with sufficient resilience to undergo this intensive treatment. These registry data lay a fundament on setting up prospective clinical controlled trials for HBOT confirming the results and investigating dose-response and most likely responders.

## Data Availability

All data produced in the present study are available upon reasonable request to the authors

## Notes

### Competing Interest Statement

The authors have declared no competing interest.

### Clinical Trial

NCT06159309

### Funding Statement

This study did not receive any funding

### Author Declarations

Medical Ethical Review Board (METC) of Erasmus University of Rotterdam gave ethical approval for this

