## Appendix - Supplementary Information for "Hyperbaric Oxygen Therapy for Long COVID: 3-Month Follow up Results from a Prospective Registry of 232 patients"

#### Figure A. Long COVID symptom severity at baseline

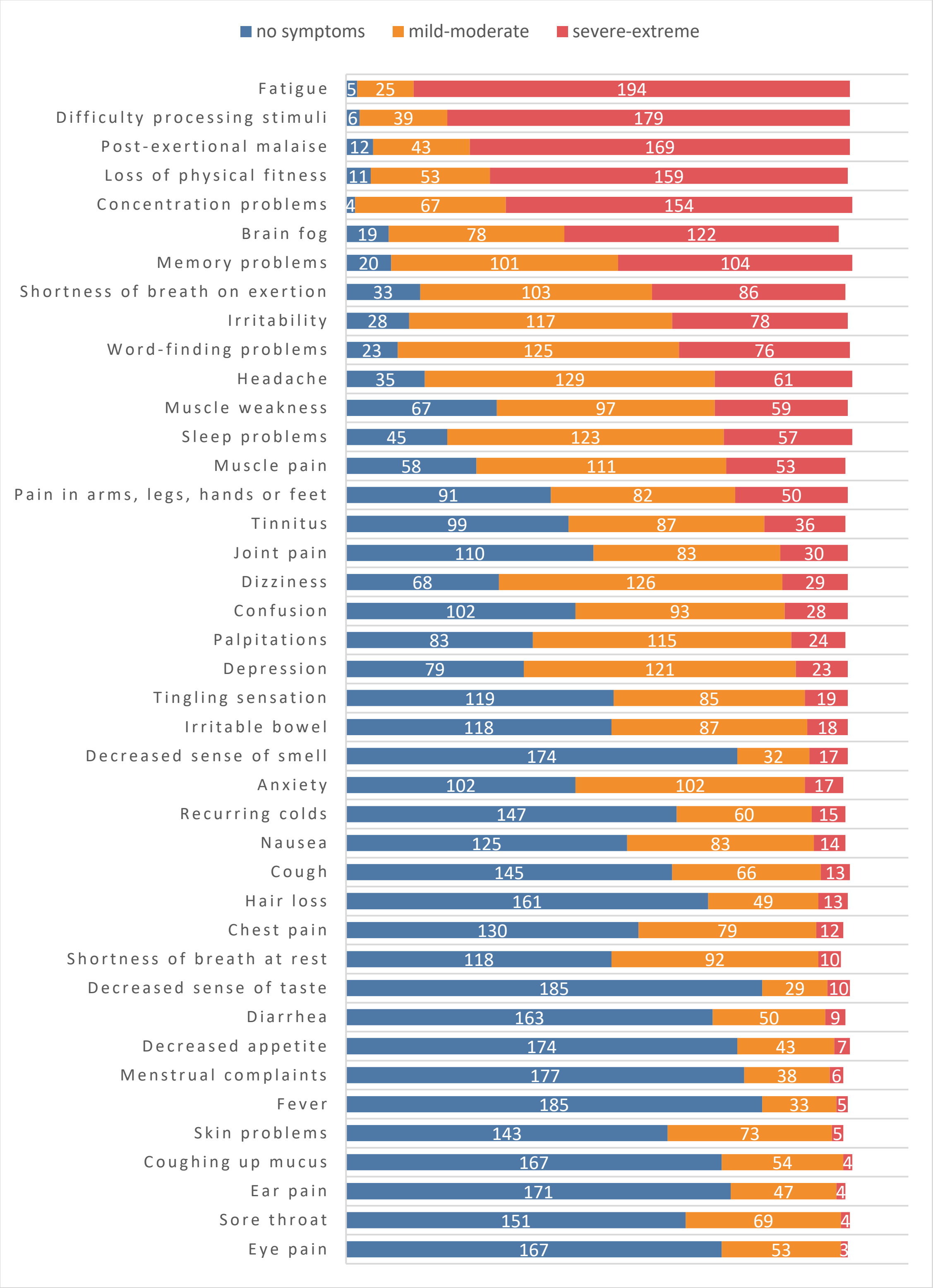


Note: Number of patients for each symptom category. Symptom severity was rated by patients on a 5-point Likert scale, and further subcategorized into; no symptoms, mild-moderate, severe-extreme.

### Table A. SF-36[i] scores

| | Baseline | Directly after HBOT | p-value* | 3-month follow up | p-value* |
|---|---|---|---|---|---|
| **MCS** | 45,5 (28,7 - 56,6) | 52,5 (38,3 - 63,3) | <0,001 | 57,0 (46,2 - 71,5) | <0,001 |
| Role limitations due to emotional problems | 100,0 (0,0 - 100,0) | 100,0 (33,3 - 100,0) | 0,059 | 100,0 (100,0 - 100,0) | <0,001 |
| Energy/fatigue | 20,0 (10,0 - 30,0) | 30,0 (15,0 – 45,0) | <0,001 | 35,0 (20,0 - 55,0) | <0,001 |
| Emotional well-being | 64,0 (52,0 - 76,0) | 70,0 (60,0 - 80,0) | <0,001 | 72,0 (60,0 - 84,0) | <0,001 |
| Social functioning | 12,5 (0,0 – 37,5) | 25,0 (0,0 – 50,0) | <0,001 | 37,5 (12,5 - 62,5) | <0,001 |
| **PCS** | 36,9 (27,2 – 45,0) | 40,6 (29,1 - 53,1) | 0,001 | 42,5 (32,2 – 52,5) | <0,001 |
| General health | 30,0 (20,0 - 40,0) | 35,0 (25,0 - 45,0) | 0,008 | 35,0 (25,0 - 50,0) | 0,031 |
| Physical functioning | 50,0 (35,0 - 70,0) | 65,0 (40,0 - 80,0) | <0,001 | 65,0 (40,0 - 80,0) | <0,001 |
| Role limitations due to physical health | 0,0 (0,0 - 0,0) | 0,0 (0,0 - 0,0) | <0,001 | 0,0 (0,0 - 0,0) | <0,001 |
| Pain | 67,5 (32,5 - 80,0) | 67,5 (45,0 - 90,0) | 0,009 | 67,5 (45,0 – 90,0) | 0,007 |

Note: Variables are shown as median (IQR).

Abbreviations: SF-36, 36-Item Short Form Survey. MCS, mental component score (range 0-100), PCS, physical component score. IQR, interquartile range

[i]SF-36 measures quality-of-life. The MCS quality-of-life on four health domains: role limitations due to emotional problems, energy/fatigue, emotional well-being and social functioning. The PCS includes general health, physical functioning, role limitations due to physical health and pain. Scores range from 0 to 100, with higher scores indicating higher quality of life.

* statistical significance as compared to baseline

**Table B. Symptom scores at baseline and 3-month follow up.**

**Anxiety symptom score**

| Baseline | 3-month follow up | |
|---|---|---|
| No symptoms (N=82) | No symptoms | 70 |
| | Mild - Moderate | 11 |
| | Severe- Extreme | 1 |
| Mild – Moderate (N=83) | No symptoms | 41 |
| | Mild - Moderate | 41 |
| | Severe- Extreme | 1 |
| Severe- Extreme (N=13) | No symptoms | 4 |
| | Mild - Moderate | 6 |
| | Severe- Extreme | 3 |

**Concentration problems symptom score**

| baseline | 3-month follow up | |
|---|---|---|
| No symptoms (N=4) | No symptoms | 3 |
| | Mild - Moderate | 1 |
| | Severe- Extreme | 0 |
| Mild – Moderate (N=50) | No symptoms | 5 |
| | Mild - Moderate | 40 |
| | Severe- Extreme | 5 |
| Severe- Extreme (N=126) | No symptoms | 7 |
| | Mild - Moderate | 70 |
| | Severe- Extreme | 49 |

**Loss of physical fitness symptom score**

| baseline | 3-month follow up | |
|---|---|---|
| No symptoms (N=10) | No symptoms | 6 |
| | Mild - Moderate | 3 |
| | Severe- Extreme | 1 |
| Mild – Moderate (N=43) | No symptoms | 7 |
| | Mild - Moderate | 22 |
| | Severe- Extreme | 14 |
| Severe- Extreme (N=123) | No symptoms | 7 |
| | Mild - Moderate | 35 |
| | Severe- Extreme | 81 |

**Diarrhea symptom score**

| baseline | 3-month follow up | |
|---|---|---|
| No symptoms (N=126) | No symptoms | 109 |
| | Mild - Moderate | 16 |
| | Severe- Extreme | 1 |
| Mild – Moderate (N=47) | No symptoms | 24 |
| | Mild - Moderate | 20 |
| | Severe- Extreme | 3 |
| Severe- Extreme (N=6) | No symptoms | 3 |
| | Mild - Moderate | 2 |
| | Severe- Extreme | 1 |

**Dizziness symptom score**

| baseline | 3-month follow up | |
|---|---|---|
| No symptoms (N=51) | No symptoms | 38 |
| | Mild - Moderate | 13 |
| | Severe- Extreme | 0 |
| Mild – Moderate (N=104) | No symptoms | 38 |
| | Mild - Moderate | 59 |
| | Severe- Extreme | 7 |
| Severe- Extreme (N=23) | No symptoms | 2 |
| | Mild - Moderate | 13 |
| | Severe- Extreme | 8 |

**Memory problems symptom score**

| baseline | 3-month follow up | |
|---|---|---|
| No symptoms (N=12) | No symptoms | 8 |
| | Mild - Moderate | 4 |
| | Severe- Extreme | 0 |
| Mild – Moderate (N=81) | No symptoms | 12 |
| | Mild - Moderate | 68 |
| | Severe- Extreme | 1 |
| Severe- Extreme (N=88) | No symptoms | 8 |
| | Mild - Moderate | 54 |
| | Severe- Extreme | 26 |

**Joint pain symptom score**

| baseline | 3-month follow up | |
|---|---|---|
| No symptoms (N=94) | No symptoms | 76 |
| | Mild - Moderate | 18 |
| | Severe- Extreme | 0 |
| Mild – Moderate (N=63) | No symptoms | 15 |
| | Mild - Moderate | 43 |
| | Severe- Extreme | 5 |
| Severe- Extreme (N=22) | No symptoms | 2 |
| | Mild - Moderate | 10 |
| | Severe- Extreme | 10 |

**Hair loss symptom score**

| baseline | 3-month follow up | |
|---|---|---|
| No symptoms (N=128) | No symptoms | 115 |
| | Mild - Moderate | 13 |
| | Severe- Extreme | 0 |
| Mild – Moderate (N=39) | No symptoms | 14 |
| | Mild - Moderate | 23 |
| | Severe- Extreme | 2 |
| Severe- Extreme (N=12) | No symptoms | 3 |
| | Mild - Moderate | 9 |
| | Severe- Extreme | 0 |

| **Palpitations symptom score** | | |
|---|---|---|
| **baseline** | **3-month follow up** | |
| No symptoms (N=70) | No symptoms | 56 |
| | Mild - Moderate | 14 |
| | Severe- Extreme | 0 |
| Mild – Moderate (N=87) | No symptoms | 27 |
| | Mild - Moderate | 54 |
| | Severe- Extreme | 6 |
| Severe- Extreme (N=21) | No symptoms | 5 |
| | Mild - Moderate | 11 |
| | Severe- Extreme | 5 |

| **Brain fog symptom score** | | |
|---|---|---|
| **baseline** | **3-month follow up** | |
| No symptoms (N=12) | No symptoms | 9 |
| | Mild - Moderate | 3 |
| | Severe- Extreme | 0 |
| Mild – Moderate (N=62) | No symptoms | 17 |
| | Mild - Moderate | 40 |
| | Severe- Extreme | 5 |
| Severe- Extreme (N=103) | No symptoms | 14 |
| | Mild - Moderate | 56 |
| | Severe- Extreme | 33 |

| **Cough symptom score** | | |
|---|---|---|
| **baseline** | **3-month follow up** | |
| No symptoms (N=120) | No symptoms | 101 |
| | Mild - Moderate | 16 |
| | Severe- Extreme | 3 |
| Mild – Moderate (N=52) | No symptoms | 27 |
| | Mild - Moderate | 22 |
| | Severe- Extreme | 3 |
| Severe- Extreme (N=7) | No symptoms | 5 |
| | Mild - Moderate | 2 |
| | Severe- Extreme | 0 |

| **Headache symptom score** | | |
|---|---|---|
| **baseline** | **3-month follow up** | |
| No symptoms (N=28) | No symptoms | 17 |
| | Mild - Moderate | 10 |
| | Severe- Extreme | 1 |
| Mild – Moderate (N=108) | No symptoms | 29 |
| | Mild - Moderate | 69 |
| | Severe- Extreme | 10 |
| Severe- Extreme (N=43) | No symptoms | 7 |
| | Mild - Moderate | 20 |
| | Severe- Extreme | 16 |

| **Skin problems symptom score** | | |
|---|---|---|
| **baseline** | **3-month follow up** | |
| No symptoms (N=112) | No symptoms | 97 |
| | Mild - Moderate | 13 |
| | Severe- Extreme | 2 |
| Mild – Moderate (N=64) | No symptoms | 27 |
| | Mild - Moderate | 35 |
| | Severe- Extreme | 2 |
| Severe- Extreme (N=3) | No symptoms | 1 |
| | Mild - Moderate | 0 |
| | Severe- Extreme | 2 |

| **Sore throat symptom score** | | |
|---|---|---|
| **baseline** | **3-month follow up** | |
| No symptoms (N=123) | No symptoms | 106 |
| | Mild - Moderate | 16 |
| | Severe- Extreme | 1 |
| Mild – Moderate (N=52) | No symptoms | 32 |
| | Mild - Moderate | 19 |
| | Severe- Extreme | 1 |
| Severe- Extreme (N=2) | No symptoms | 2 |
| | Mild - Moderate | 0 |
| | Severe- Extreme | 0 |

| **Fever symptom score** | | |
|---|---|---|
| **baseline** | **3-month follow up** | |
| No symptoms (N=154) | No symptoms | 144 |
| | Mild - Moderate | 9 |
| | Severe- Extreme | 1 |
| Mild – Moderate (N=20) | No symptoms | 10 |
| | Mild - Moderate | 9 |
| | Severe- Extreme | 1 |
| Severe- Extreme (N=4) | No symptoms | 1 |
| | Mild - Moderate | 2 |
| | Severe- Extreme | 1 |

| **Shortness of breath on exertion symptom score** | | |
|---|---|---|
| **baseline** | **3-month follow up** | |
| No symptoms (N=26) | No symptoms | 19 |
| | Mild - Moderate | 7 |
| | Severe- Extreme | 0 |
| Mild – Moderate (N=85) | No symptoms | 16 |
| | Mild - Moderate | 63 |
| | Severe- Extreme | 6 |
| Severe- Extreme (N=68) | No symptoms | 5 |
| | Mild - Moderate | 33 |
| | Severe- Extreme | 30 |

| **Shortness of breath at rest symptom score** | | |
|---|---|---|
| **baseline** | **3-month follow up** | |
| No symptoms (N=102) | No symptoms | 86 |
| | Mild - Moderate | 16 |
| | Severe- Extreme | 0 |
| Mild – Moderate (N=69) | No symptoms | 27 |
| | Mild - Moderate | 40 |
| | Severe- Extreme | 2 |
| Severe- Extreme (N=5) | No symptoms | 2 |
| | Mild - Moderate | 2 |
| | Severe- Extreme | 1 |

| **Menstrual complaints symptom score (F)** | | |
|---|---|---|
| **baseline** | **3-month follow up** | |
| No symptoms (N=90) | No symptoms | 78 |
| | Mild - Moderate | 8 |
| | Severe- Extreme | 4 |
| Mild – Moderate (N=31) | No symptoms | 10 |
| | Mild - Moderate | 17 |
| | Severe- Extreme | 4 |
| Severe- Extreme (N=4) | No symptoms | 1 |
| | Mild - Moderate | 1 |
| | Severe- Extreme | 2 |

| **Nausea symptom score** | | |
|---|---|---|
| **baseline** | **3-month follow up** | |
| No symptoms (N=104) | No symptoms | 85 |
| | Mild - Moderate | 18 |
| | Severe- Extreme | 1 |
| Mild – Moderate (N=66) | No symptoms | 28 |
| | Mild - Moderate | 34 |
| | Severe- Extreme | 4 |
| Severe- Extreme (N=9) | No symptoms | 2 |
| | Mild - Moderate | 5 |
| | Severe- Extreme | 2 |

| **Difficulty processing stimuli symptom score** | | |
|---|---|---|
| **baseline** | **3-month follow up** | |
| No symptoms (N=6) | No symptoms | 3 |
| | Mild - Moderate | 1 |
| | Severe- Extreme | 2 |
| Mild – Moderate (N=27) | No symptoms | 3 |
| | Mild - Moderate | 20 |
| | Severe- Extreme | 4 |
| Severe- Extreme (N=146) | No symptoms | 8 |
| | Mild - Moderate | 73 |
| | Severe- Extreme | 65 |

| **Eye pain symptom score** | | |
|---|---|---|
| **baseline** | **3-month follow up** | |
| No symptoms (N=132) | No symptoms | 116 |
| | Mild - Moderate | 15 |
| | Severe- Extreme | 1 |
| Mild – Moderate (N=46) | No symptoms | 28 |
| | Mild - Moderate | 16 |
| | Severe- Extreme | 2 |
| Severe- Extreme (N=1) | No symptoms | 1 |
| | Mild - Moderate | 0 |
| | Severe- Extreme | 0 |

| **Ear pain symptom score** | | |
|---|---|---|
| **baseline** | **3-month follow up** | |
| No symptoms (N=139) | No symptoms | 133 |
| | Mild - Moderate | 5 |
| | Severe- Extreme | 1 |
| Mild – Moderate (N=37) | No symptoms | 24 |
| | Mild - Moderate | 13 |
| | Severe- Extreme | 0 |
| Severe- Extreme (N=2) | No symptoms | 1 |
| | Mild - Moderate | 0 |
| | Severe- Extreme | 1 |

| **Tinnitus symptom score** | | |
|---|---|---|
| **baseline** | **3-month follow up** | |
| No symptoms (N=77) | No symptoms | 66 |
| | Mild - Moderate | 10 |
| | Severe- Extreme | 1 |
| Mild – Moderate (N=74) | No symptoms | 21 |
| | Mild - Moderate | 42 |
| | Severe- Extreme | 11 |
| Severe- Extreme (N=26) | No symptoms | 2 |
| | Mild - Moderate | 8 |
| | Severe- Extreme | 16 |

| **Pain in arms, legs, hands or feet symptom score** | | |
|---|---|---|
| **baseline** | **3-month follow up** | |
| No symptoms (N=74) | No symptoms | 50 |
| | Mild - Moderate | 22 |
| | Severe- Extreme | 2 |
| Mild – Moderate (N=67) | No symptoms | 26 |
| | Mild - Moderate | 35 |
| | Severe- Extreme | 6 |
| Severe- Extreme (N=39) | No symptoms | 3 |
| | Mild - Moderate | 20 |
| | Severe- Extreme | 16 |

| Chest pain symptom score | | |
|---|---|---|
| baseline | 3-month follow up | |
| No symptoms (N=110) | No symptoms | 103 |
| | Mild - Moderate | 7 |
| | Severe- Extreme | 0 |
| Mild – Moderate (N=62) | No symptoms | 24 |
| | Mild - Moderate | 36 |
| | Severe- Extreme | 2 |
| Severe- Extreme (N=7) | No symptoms | 0 |
| | Mild - Moderate | 5 |
| | Severe- Extreme | 2 |

| Irritability symptom score | | |
|---|---|---|
| baseline | 3-month follow up | |
| No symptoms (N=23) | No symptoms | 17 |
| | Mild - Moderate | 6 |
| | Severe- Extreme | 0 |
| Mild – Moderate (N=95) | No symptoms | 15 |
| | Mild - Moderate | 70 |
| | Severe- Extreme | 10 |
| Severe- Extreme (N=61) | No symptoms | 6 |
| | Mild - Moderate | 38 |
| | Severe- Extreme | 17 |

| Irritable bowel symptom score | | |
|---|---|---|
| baseline | 3-month follow up | |
| No symptoms (N=98) | No symptoms | 87 |
| | Mild - Moderate | 10 |
| | Severe- Extreme | 1 |
| Mild – Moderate (N=68) | No symptoms | 28 |
| | Mild - Moderate | 38 |
| | Severe- Extreme | 2 |
| Severe- Extreme (N=13) | No symptoms | 1 |
| | Mild - Moderate | 5 |
| | Severe- Extreme | 7 |

| Sleep problems symptom score | | |
|---|---|---|
| baseline | 3-month follow up | |
| No symptoms (N=38) | No symptoms | 22 |
| | Mild - Moderate | 15 |
| | Severe- Extreme | 1 |
| Mild – Moderate (N=100) | No symptoms | 23 |
| | Mild - Moderate | 63 |
| | Severe- Extreme | 14 |
| Severe- Extreme (N=42) | No symptoms | 2 |
| | Mild - Moderate | 20 |
| | Severe- Extreme | 20 |

| Coughing up mucus symptom score | | |
|---|---|---|
| baseline | 3-month follow up | |
| No symptoms (N=136) | No symptoms | 118 |
| | Mild - Moderate | 17 |
| | Severe- Extreme | 1 |
| Mild – Moderate (N=42) | No symptoms | 29 |
| | Mild - Moderate | 12 |
| | Severe- Extreme | 1 |
| Severe- Extreme (N=3) | No symptoms | 2 |
| | Mild - Moderate | 0 |
| | Severe- Extreme | 1 |

| Depression symptom score | | |
|---|---|---|
| baseline | 3-month follow up | |
| No symptoms (N=61) | No symptoms | 46 |
| | Mild - Moderate | 15 |
| | Severe- Extreme | 0 |
| Mild – Moderate (N=103) | No symptoms | 42 |
| | Mild - Moderate | 59 |
| | Severe- Extreme | 2 |
| Severe- Extreme (N=15) | No symptoms | 4 |
| | Mild - Moderate | 7 |
| | Severe- Extreme | 4 |

| Muscle pain symptom score | | |
|---|---|---|
| baseline | 3-month follow up | |
| No symptoms (N=51) | No symptoms | 36 |
| | Mild - Moderate | 12 |
| | Severe- Extreme | 3 |
| Mild – Moderate (N=90) | No symptoms | 20 |
| | Mild - Moderate | 57 |
| | Severe- Extreme | 13 |
| Severe- Extreme (N=36) | No symptoms | 5 |
| | Mild - Moderate | 12 |
| | Severe- Extreme | 19 |

| Muscle weakness symptom score | | |
|---|---|---|
| baseline | 3-month follow up | |
| No symptoms (N=54) | No symptoms | 36 |
| | Mild - Moderate | 17 |
| | Severe- Extreme | 1 |
| Mild – Moderate (N=78) | No symptoms | 19 |
| | Mild - Moderate | 48 |
| | Severe- Extreme | 11 |
| Severe- Extreme (N=45) | No symptoms | 3 |
| | Mild - Moderate | 19 |
| | Severe- Extreme | 23 |

| Recurring colds symptom score | | |
|---|---|---|
| baseline | 3-month follow up | |
| No symptoms (N=118) | No symptoms | 97 |
| | Mild - Moderate | 20 |
| | Severe- Extreme | 1 |
| Mild – Moderate (N=49) | No symptoms | 25 |
| | Mild - Moderate | 23 |
| | Severe- Extreme | 1 |
| Severe- Extreme (N=10) | No symptoms | 2 |
| | Mild - Moderate | 6 |
| | Severe- Extreme | 2 |

| Tingling sensation symptom score | | |
|---|---|---|
| baseline | 3-month follow up | |
| No symptoms (N=97) | No symptoms | 81 |
| | Mild - Moderate | 15 |
| | Severe- Extreme | 1 |
| Mild – Moderate (N=70) | No symptoms | 32 |
| | Mild - Moderate | 34 |
| | Severe- Extreme | 4 |
| Severe- Extreme (N=13) | No symptoms | 3 |
| | Mild - Moderate | 5 |
| | Severe- Extreme | 5 |

| Post-exertional malaise symptom score | | |
|---|---|---|
| baseline | 3-month follow up | |
| No symptoms (N=6) | No symptoms | 5 |
| | Mild - Moderate | 1 |
| | Severe- Extreme | 0 |
| Mild – Moderate (N=38) | No symptoms | 10 |
| | Mild - Moderate | 22 |
| | Severe- Extreme | 6 |
| Severe- Extreme (N=136) | No symptoms | 9 |
| | Mild - Moderate | 36 |
| | Severe- Extreme | 91 |

| Fatigue symptom score | | |
|---|---|---|
| baseline | 3-month follow up | |
| No symptoms (N=2) | No symptoms | 1 |
| | Mild - Moderate | 0 |
| | Severe- Extreme | 1 |
| Mild – Moderate (N=21) | No symptoms | 0 |
| | Mild - Moderate | 13 |
| | Severe- Extreme | 8 |
| Severe- Extreme (N=155) | No symptoms | 5 |
| | Mild - Moderate | 50 |
| | Severe- Extreme | 100 |

| Decreased sense if smell symptom score | | |
|---|---|---|
| baseline | 3-month follow up | |
| No symptoms (N=142) | No symptoms | 140 |
| | Mild - Moderate | 1 |
| | Severe- Extreme | 1 |
| Mild – Moderate (N=28) | No symptoms | 15 |
| | Mild - Moderate | 11 |
| | Severe- Extreme | 2 |
| Severe- Extreme (N=9) | No symptoms | 0 |
| | Mild - Moderate | 4 |
| | Severe- Extreme | 5 |

| Decreased sense of taste score | | |
|---|---|---|
| baseline | 3-month follow up | |
| No symptoms (N=152) | No symptoms | 147 |
| | Mild - Moderate | 5 |
| | Severe- Extreme | 0 |
| Mild – Moderate (N=21) | No symptoms | 12 |
| | Mild - Moderate | 8 |
| | Severe- Extreme | 1 |
| Severe- Extreme (N=7) | No symptoms | 1 |
| | Mild - Moderate | 2 |
| | Severe- Extreme | 4 |

| Decreased appetite symptom score | | |
|---|---|---|
| baseline | 3-month follow up | |
| No symptoms (N=140) | No symptoms | 127 |
| | Mild - Moderate | 12 |
| | Severe- Extreme | 1 |
| Mild – Moderate (N=35) | No symptoms | 17 |
| | Mild - Moderate | 17 |
| | Severe- Extreme | 1 |
| Severe- Extreme (N=4) | No symptoms | 1 |
| | Mild - Moderate | 1 |
| | Severe- Extreme | 2 |

| Confusion symptom score | | |
|---|---|---|
| baseline | 3-month follow up | |
| No symptoms (N=82) | No symptoms | 71 |
| | Mild - Moderate | 11 |
| | Severe- Extreme | 0 |
| Mild – Moderate (N=79) | No symptoms | 39 |
| | Mild - Modera te | 36 |
| | Severe- Extreme | 4 |
| Severe- Extreme (N=19) | No symptoms | 5 |
| | Mild - Moderate | 11 |
| | Severe- Extreme | 3 |

| Word-finding problems symptom score | | |
|---|---|---|
| **baseline** | **3-month follow up** | |
| No symptoms (N=17) | No symptoms | 16 |
| | Mild - Moderate | 1 |
| | Severe- Extreme | 0 |
| Mild – Moderate (N=103) | No symptoms | 24 |
| | Mild - Moderate | 77 |
| | Severe- Extreme | 2 |
| Severe- Extreme (N=59) | No symptoms | 4 |
| | Mild - Moderate | 39 |
| | Severe- Extreme | 16 |

Note: Number of patients for each symptom category. Symptom severity was rated by patients on a 5-point Likert scale, and further subcategorized into; no symptoms, mild-moderate, severe-extreme.
